# Urinary bisphenol A exposure in relation to liver function abnormalities among U.S. adults: results from the National Health and Nutrition Examination Survey, 2003-2016

**DOI:** 10.1101/2023.10.01.23296398

**Authors:** Tiantian Zhang, Guang Huang, Tongshuai Wang, Jie Chen, Xiangyu Zhou, Wenming Shi

**Affiliations:** School of Public Health, Fudan University, Shanghai, 200032, China; School of Public Health, Bengbu Medical College, Bengbu, Anhui, China; Hongqiao International Institute of Medicine, Shanghai Tongren Hospital, School of Medicine of Shanghai Jiaotong University, Shanghai 20050, China; Department of Big Data in Health Science School of Public Health, Zhejiang University School of Medicine, Hangzhou, Zhejiang, China; Obstetrics and Gynecology Hospital of Fudan University, State Key Lab of Genetic Engineering, Fudan University Shanghai Medical College, Shanghai 200011,China; School of Public Health, Li Ka Shing Faculty of Medicine, The University of Hong Kong, Hong Kong Special Administrative Region, Hong Kong, China

**Author notes:** Correspondence: Wenming Shi, PhD, Address: No. 130, Dong’an Road, Xuhui District, Shanghai 200032, China. These two authors contributed equally to this work (Tiantian Zhang, Guang Huang).

**Keywords:** Bisphenol A, Liver injury, Liver enzymes, NHANES, Total bilirubin

## Abstract

**Background:** Bisphenol A (BPA) is an endocrine-disrupting chemical that has been linked with various health outcomes. However, few studies using nationally representative data have assessed the association between BPA exposure with liver function. In addition, whether behavior characteristics of smoking and alcohol use modify such association has been underexplored.

**Methods:** Using data from seven cycles of the National Health and Nutrition Examination Surveys (NHANES) among 11,750 adults from 2003 to 2016, we investigated the relationship between urinary BPA with liver function indicators. BPA concentration in urine was measured by using high performance liquid chromatography with atmospheric pressure chemical ionization–mass spectrometry.

We estimated BPA concentration after controlling for creatinine and normalized the asymmetrical distribution using natural logarithmic transformation (ln-BPA/Cr). Eight liver function indicators in serum were examined. Multivariate linear regression models were performed to explore the association between urinary BPA with changes in liver indicators. Stratified analyses examined whether these associations varied by sex, age, smoking, or drinking.

**Findings:** Of the 11,750 participants (49.5% men), the mean age was 43.9 years. Per unit increment in ln-BPA/Cr was positively related to alkaline phosphotase (ALP) and total bilirubin (TBIL), while inversely with albumin (ALB). In quartile analysis, the highest BPA exposure was associated with higher aspartate aminotransferase (AST), ALP, and TBIL, while with decreased ALB (all P_trend_ < 0.01). These associations for specific liver indicators (i.e., ALT, AST, Gamma-glutamyl transferase [GGT], and TBIL) were stronger in smokers and alcohol drinkers. Moreover, we found that BPA exposure with higher ALP in older adults (≥ 60 years) but no sex difference.

**Interpretation:** This nationally representative study suggested that urinary BPA was associated with elevated AST, ALP, TBIL, and inversely with ALB among U.S. adults. The associations were more evident in smokers and drinkers but no sex difference. Better understanding of the mechanisms is needed for improving liver and planetary health.

**Funding:** National Natural Science Foundation of China.

## Introduction

Over the past decades, liver diseases have become one of the leading causes of mortality and illness globally (1). Studies have reported that more than 800 million people suffer from chronic liver disease worldwide, with two million deaths per year (1,2). People with advanced chronic liver disease have high rates of hospital admission and generally need more healthcare services that have a substantial economic cost (3). Therefore, it is imperative to work together to understand the risk factors of liver health to better cope with the rising tide of liver disease. Liver function testing (LFT) is one of the commonly used clinical assessments for the diagnosis of liver injury (4). In recent years, increasing evidence has suggested that many chemicals and environmental pollutants can lead to liver dysfunction and increase liver disease risk (5,6).

Bisphenol A (BPA) is a phenolic compound widely used in the manufacture of polycarbonate plastics and epoxy resin. With the rapidly increasing production of BPA, it is reported that the production is projected to be about 10.6 million tons in 2022 (7). Besides the environmental substrates, BPA can also be found in food and beverage containers, bottles and cans, tap water pipes, toys, electronic devices, and even in the composites in dental restorations (8). Thus, people are continuously exposed to BPA through ingestion, inhalation, dermal contact, and other pathways (9). A growing number of studies indicate that BPA is an endocrine-disrupting chemical, which is related to multiple adverse health outcomes including cardiovascular diseases (CVD) (10), diabetes (11), kidney disease (12), and reproductive health (13,14). Animal experiments showed that BPA-treated rats had elevated levels of alanine aminotransferase (ALT) and aspartate aminotransferase (AST) as well as marked defects in liver morphology (15). A panel study conducted in Korea presented that urinary BPA was associated with ALT, AST, and Gamma glutamyl transferase (GGT) in the elderly (16). However, previous studies are performed with several limitations of analyzing limited liver function indicators, or in a special population (16,17), the knowledge about BPA exposure on other liver function parameters such as total bilirubin (TBIL), albumin (ALB), and total protein (TP) is under-examined. Evidence has suggested that TBIL is a critical indicator for the diagnosis of liver disease, and serum TP and ALB levels can indicate chronic liver injury and hepatocyte reserve function (4,18). Therefore, using a general population study to comprehensively evaluate the impacts of BPA on liver function indicators is merited. In addition, considering smoking and alcohol consumption are prevalent behavior factors around the world, however, whether these behavior characteristics modified the association between BPA and liver function remains unclear.

Given the high-volume industrial production of BPA worldwide, identifying the effects of BPA exposure on liver function is critical for improving liver and planetary health. Using a nationally population-representative study in the United States, we aimed to investigate the association between urinary BPA with multiple liver function indicators among U.S. adults. Moreover, we evaluated whether the associations varied by sex, age, smoking, and alcohol drinking.

## Methods

### Study design and participants

The National Health and Nutrition Examination Survey (NHANES) is an ongoing national-representative survey of the U.S. non-institutionalized population. Detailed study design and study population have been described at https://www.cdc.gov/nchs/nhanes/index.html. Briefly, a stratified, multistage probability sampling was applied to collect health and nutritional status of adults and children in the U.S. annually by the National Center for Health Statistics (NCHS) (https://wwwn.cdc.gov/nchs/nhanes/default.aspx). NCHS-trained professionals conducted interviews in participants’ homes, whereas extensive physical measurements, including blood and urine collection, were conducted at mobile exam centers.

In this study, we combined data from seven cycles from NHANES (2003-2004, 2005-2006, 2007-2008, 2009-2010, 2011-2012, 2013-2014, and 2015-2016) with available individual BPA measurement. Of the 71,058 participants in total, we excluded individuals aged less than 18 years old at the time of participation (n=29,151), with missing data of BPA (n=26,480), without urinary creatinine and health outcomes (n=3,677), resulting in 11,750 participants included in the final analyses. The NHANES study was approved by the ethical review committee of the NCHS. All participants provided written informed consent. Figure S1 shows the selection process of the participants. This study followed the Strengthening the Reporting of Observational Studies in Epidemiology (STROBE) reporting guidelines (19).

### Measurement of urinary BPA

As part of NHANES ongoing biomonitoring program, spot urine samples were collected from one-third random subsample of the participants. The urine samples were frozen and stored on dry ice for shipping to the U.S. National Center for Environmental Health before testing. Urinary BPA concentration was measured by using solid phase extraction coupled with high performance liquid chromatography with atmospheric pressure chemical ionization–mass spectrometry (HPLC/MS-MS). To control for dilution, the creatinine-corrected BPA concentration (BPA/Cr, ng/mg) was applied in the current study. The limit of detection (LOD) of urinary BPA values was in the level of 0.4 ng/ml for each cycle in NHANES. To reduce the bias from the relatively high rate of individuals (around 10%) with BPA concentrations below the LOD, we excluded them from our analysis.

### Liver function tests

ALT and AST are liver enzyme parameters released after hepatocytes necrosis or liver cell membrane injury, they are elevated during acute liver damage (4). ALT/AST is the ratio of ALT and AST concentration, which is a sensitivity indicator of liver injury (20). ALP is a marker of cholestasis, and GGT is used in clinical practice to ascertain the liver source of an elevated ALP (21). Serum TBIL level is affected by the balance between bilirubin production and clearance, and hepatocytes are the main site of bilirubin clearance (22). Elevated serum bilirubin levels usually suggest liver damage or obstruction of the bile duct. Decreased ALB or TP mainly implies limited liver synthesis (4).

In this study, serum liver function parameters were measured by different methods using Beckman Coulter technique. During 2003-2006, serum liver indicators were tested on a Beckman Synchron LX20 using an enzymatic rate method. In 2007-2016, the levels of serum liver function indicators were measured on a Beckman Unicel DxC800 Synchron using a kinetic rate method (23). The ALT/AST (DeRitis) ratio was calculated accordingly.

### Covariates

We collected the covariates or potential confounding that are linked to liver function or urinary BPA according to common sense or previous findings(16,17). The covariates included demographic characteristics of age, sex, body mass index (BMI), educational level (less than high school, high school, some college and more), race/ethnicity, poverty income ratio (PIR), smoking status (current, former, never), alcohol drinking (yes or no), and main chronic conditions including hypertension, diabetes, and cardiovascular disease (CVD), which were obtained using validated questionnaire during the family interview. Race/ ethnicity was categorized as “Mexican American”, “other Hispanic”, “non-Hispanic white”, “non-Hispanic black”, and “other race”. PIR is a measurement of socioeconomic status and represents the ratio of household income to the poverty threshold after accounting for inflation and family size (24). Hypertension was defined as systolic blood pressure (SBP) ≥140 mmHg and/or diastolic blood pressure (DBP) ≥ 90 mmHg, and/or self-reported doctor-diagnosed hypertension, and/ or taking antihypertensive medicine. Diabetes was defined as measured fasting blood glucose ≥ 7.0 mmol/l (or oral glucose tolerance test, OGTT ≥ 11.1mmol/l), and/ or glycohemoglobin (HbA1c) ≥ 6.5%, with self-reported doctor-diagnosed diabetes, and/ or taking anti-diabetes medicine or insulin. CVD was assessed if a participant had one of the following diagnosed diseases: congestive heart failure, coronary heart disease, heart attack (or myocardial infarction), angina pectoris, and stroke.

### Statistical analyses

Following the CDC guideline (https://wwwn.cdc.gov/nchs/nhanes/tutorials/default.aspx), all analyses were performed using appropriate weights as recommended by NCHS, to generate accurate estimates nationally, adjusting for the oversampling of minority groups in the NHANES. Descriptive characteristics were summarized for the total participants included and participants in each cycle from 2003 to 2016. Continuous variable was presented as median (interquartile range, IQR), and categorized variable was expressed as absolute value and percentages. Considering the missing value for the covariates (ranging from 0.12% for educational level to 12.7% for alcohol drinking), multiple imputation (MI) procedure was performed to deal with the missing in our analysis. The trend of urinary exposure concentrations of BPA across seven cycles from 2003 to 2016 was summarized. Moreover, we evaluated creatinine-corrected BPA distributions (BPA/Cr, ng/mg) by participants’ characteristics.

The creatinine-corrected BPA was a natural logarithm transformed to normalize its distribution. Survey-weighted linear regression models were conducted to calculate the estimates of β and 95% confidence intervals (95% CI) on the association of urinary BPA with liver function indicators. We fitted three regression models: 1) model 1: crude model; model 2 adjusting for age, sex, BMI, educational level, race/ethnicity, PIR, smoking status, and alcohol drinking; 3) further adjusting for hypertension, diabetes, and CVD based on model 2. Furthermore, we divided the ln-transformed BPA concentrations (ln-BPA/Cr) into four quartiles to further evaluate the associations by using the fully adjusted model, with reference to the lowest quartile. Linear trend tests were conducted by applying quartiles as a continuous variable in the model. To explore the exposure-response relationship between urinary BPA with liver function indicators, restricted cubic spline (RCS) analysis was performed.

To examine the potential effect modifiers, we conducted several stratified analyses by the following variables: sex (male or female), age (≥ 60 years or < 60 years), smoking exposure (yes or no), and alcohol drinking (yes or no). Differences between strata was examined by z-test, the formula was expressed as below:

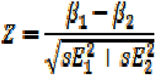

Where β_1_ and β_2_ were the effect estimates for each subgroup, SE_1_ and SE_2_ were their standard errors, respectively (25).

All statistical analyses were performed by using SAS software version 9.4 (SAS Institute Inc, Cary, NC). Two-tailed *P*-value < 0.05 was considered statistical significant.

## Results

The survey-weighted, demographic characteristics and liver function indicators of the participants were presented in Table 1. Of the 11,750 participants (49.5% men), the median age was 43.9 (IQR: 30.4 to 57.9) years. Most of the participants were non-Hispanic white (67.24%), with an educational level at some college and more, and nearly half had smoke exposure (Table 1). The median creatinine concentration was 11.70 mg/l during the overall period (Table 1), ranging from 10.7 mg/l during 2011-2012 to 12.56 mg/l during 2005-2006 (Table S1). The trend of BPA exposure concentrations in urine among the participants from 2003 to 2016 was presented in Figure 1. Overall, the urinary concentrations of BPA decreased over time except for an increment in two cycles (2005-2006 and 2009-2010) among the participants. Moreover, in the univariate analysis, we found a higher level of creatinine-corrected BPA in women and non-Hispanic White (Table S2). Table S3 presents the distribution of eight liver function indicators in serum among the participants included in each cycle. The median level of each liver indicator was similar during seven cycles in the NHANES (Table S3).

**Figure 1.**
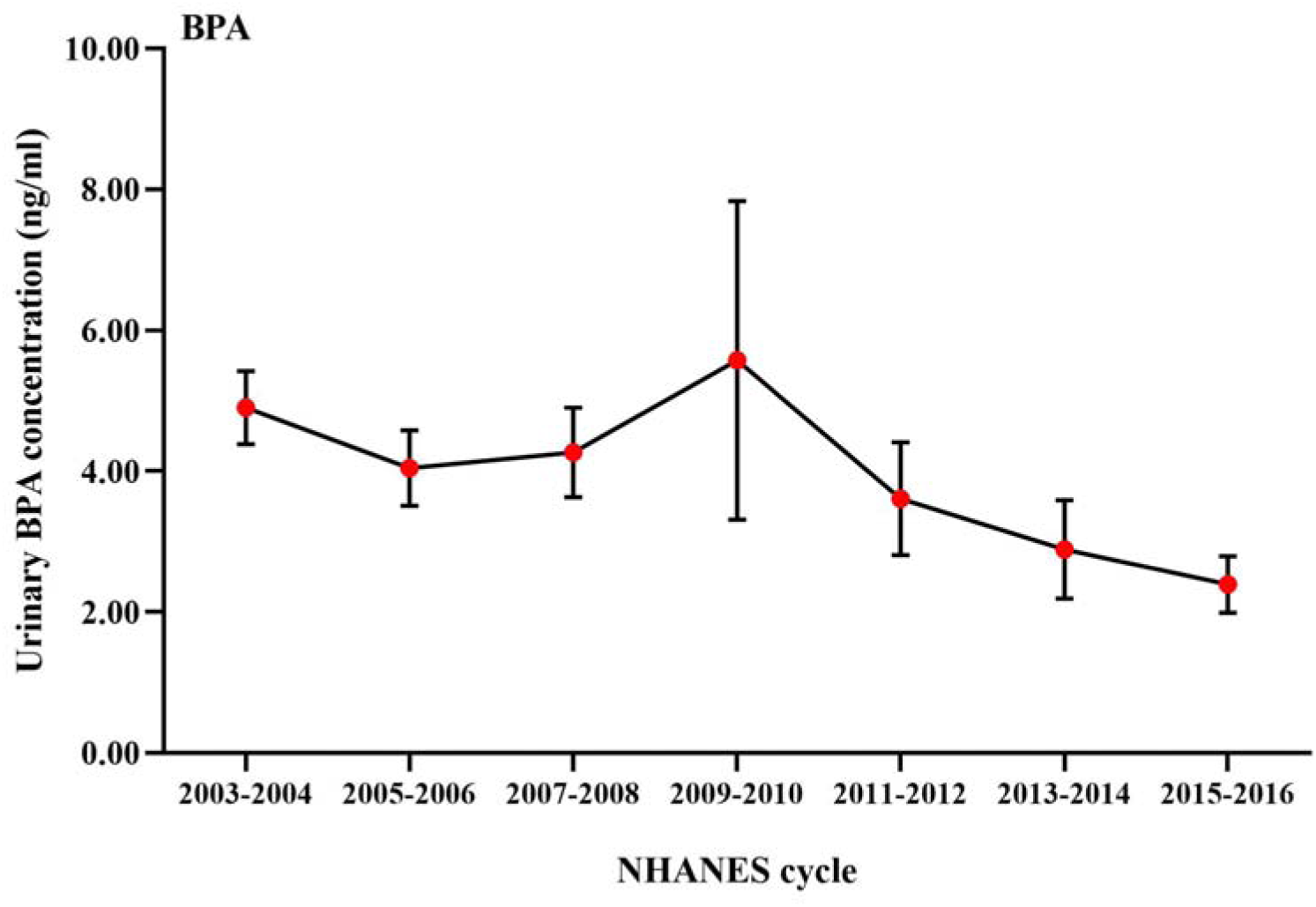
Descriptive trend of the urinary bisphenol A concentrations among U.S. adults. Note: Data are presented as mean with 95% confidence interval (95% CI), sampling weights were applied for calculation of BPA concentrations.

**Table 1.** Survey-weighted, characteristics of the participants included in the NHANES 2003-2016 (N=11,750).

| Variables | Total (n=11750) |
| --- | --- |
| Age | 43.9 (30.4, 57.9) |
| Sex |  |
| Men | 5816 (49.5) |
| Women | 5934 (50.5) |
| BMI (kg/m<sup>2</sup>) | 27.69 (23.99, 32.29) |
| Race/ethnicity |  |
| Mexican American | 1978 (8.4) |
| Other Hispanic | 1037 (5.3) |
| Non-Hispanic White | 4994 (67.2) |
| Non-Hispanic Black | 2677 (12.2) |
| Other Race | 1064 (6.9) |
| Education |  |
| Less than high school | 1217 (5.6) |
| High school | 1894 (12.0) |
| Some college and more | 8639 (82.4) |
| Poverty income ratio (PIR) | 2.82 (1.43, 4.77) |
| Smoking status |  |
| Never | 6483 (54.6) |
| Previous | 2672 (23.3) |
| Current | 2595 (22.1) |
| Alcohol consumption |  |
| No | 3418 (24.1) |
| Yes | 8332 (75.9) |
| Hypertension |  |
| No | 5414 (47.9) |
| Yes | 6336 (52.1) |
| Diabetes |  |
| No | 9766 (86.8) |
| Yes | 1984 (13.2) |
| Cardiovascular disease |  |
| No | 10538 (91.7) |
| Yes | 1212 (8.3) |
| Liver function parameters |  |
| ALT (U/L) | 20.73 (15.98, 28.57) |
| AST (U/L) | 22.59 (19.22, 27.16) |
| ALT/ AST | 0.93 (0.77, 1.14) |
| GGT (U/L) | 18.36 (12.90, 28.28) |
| ALP (U/L) | 63.23 (52.01, 76.47) |
| TBIL (mg/dL) | 0.63 (0.47, 0.85) |
| ALB (g/dL) | 4.25 (4.03, 4.47) |
| TP (g/dL) | 7.05 (6.76, 7.35) |
| Creatinine (mg/l) | 11.70 (7.10, 17.38) |

Note: Data are presented as median (IQR) or N (%); sampling weights were applied for calculation of demographic characteristics; N reflect the study sample while percentages reflect the survey-weighted. ALT: Alanine aminotransferase; AST: Aspartate aminotransferase; GGT: Gamma glutamyl transferase; ALP: Alkaline phosphatase; TBIL: Total bilirubin; ALB: albumin; TP: Total protein.

Table 2 shows the association of urinary BPA exposure with changes in liver function indicators among the participants. After controlling for confounders, we found that urinary BPA was positively related to elevated levels of ALP and TBIL, while inversely with ALB (Model 2). After further adjusting for hypertension, diabetes, and CVD in Model 3, similar associations were observed for BPA with ALP (adjusted β= 0.729, 95% CI: 0.070 to 1.389), TBIL (β=0.014, 95% CI: 0.005 to 0.023), and ALB (β= -0.011, 95% CI: -0.020 to -0.003), respectively (Table 2).

**Table 2.** Associations of urinary BPA exposure with eight liver biomarkers among the participants in NHANES 2003-2016.

| Liver<br>function<br>indicators | Model 1 |  | Model 2 |  | Model 3 |  |
| --- | --- | --- | --- | --- | --- | --- |
| | $\beta$ (95% CI) | P-value | $\beta$ (95% CI) | P-value | $\beta$ (95% CI) | P-value |
| ALT | -0.305 (-0.764 to 0.153) | 0.194 | 0.395 (-0.024 to 0.815) | 0.067 | 0.388 (-0.026 to 0.803) | 0.069 |
| AST | -0.005 (-0.313 to 0.302) | 0.973 | 0.294 (-0.011 to 0.600) | 0.061 | 0.287 (-0.014 to 0.588) | 0.065 |
| ALT/AST | -0.012 (-0.021 to -0.003) | **0.011** | 0.004 (-0.004 to 0.011) | 0.360 | 0.003 (-0.004 to 0.011) | 0.369 |
| GGT | -0.020 (-0.658 to 0.619) | 0.951 | 0.639 (-0.005 to 1.283) | 0.054 | 0.594 (-0.041 to 1.228) | 0.069 |
| ALP | 0.731 (0.059 to 1.403) | **0.035** | 0.752 (0.085 to 1.419) | **0.029** | 0.729 (0.070 to 1.389) | **0.032** |
| TBIL | 0.001 (-0.009 to 0.010) | 0.902 | 0.014 (0.005 to 0.023) | **0.003** | 0.014 (0.005 to 0.023) | **0.003** |
| ALB | -0.025 (-0.035 to -0.015) | **<0.001** | -0.012 (-0.020 to -0.003) | **0.008** | -0.011 (-0.020 to -0.003) | **0.009** |
| TP | -0.013 (-0.025 to -0.001) | **0.046** | 0.003 (-0.009 to 0.015) | 0.624 | 0.003 (-0.009 to 0.014) | 0.649 |

Note: the bold value indicated statistically significant. The estimates was obtained from per unit increment in natural logarithm-transform BPA/Cr).
Model 1: crude model;
Model 2: adjusted for age, sex, BMI, educational level, race/ethnicity, PIR, smoking status, and alcohol consumption;
Model 3: further adjusted for hypertension, diabetes, and cardiovascular disease.

Figure 2 presents the adjusted associations between urinary BPA with liver function indicators stratified by ln(BPA/Cr) quartiles. Compared with the lowest quartile (reference), the highest quartile BPA exposure was associated with AST (β=1.18, 95% CI: 0.28 to 2.08), ALP (β= 2.05, 0.74 to 3.37), TBIL (β=0.04, 0.02 to 0.07), and inversely with ALB (β=-0.03, -0.05 to -0.01). Trend analysis gave a significant association between higher BPA exposure with liver function indicators except for TP (Figure 2). The RCS plot revealed that an exposure-response relationship between ln(BPA/Cr) with an increment of ALP. Higher ln(BPA/Cr) was positively associated with ALT/AST. Moreover, we observed a decrement of TBIL related to ln (BPA/Cr) in the RCS model (Figure S2).

**Figure 2.**
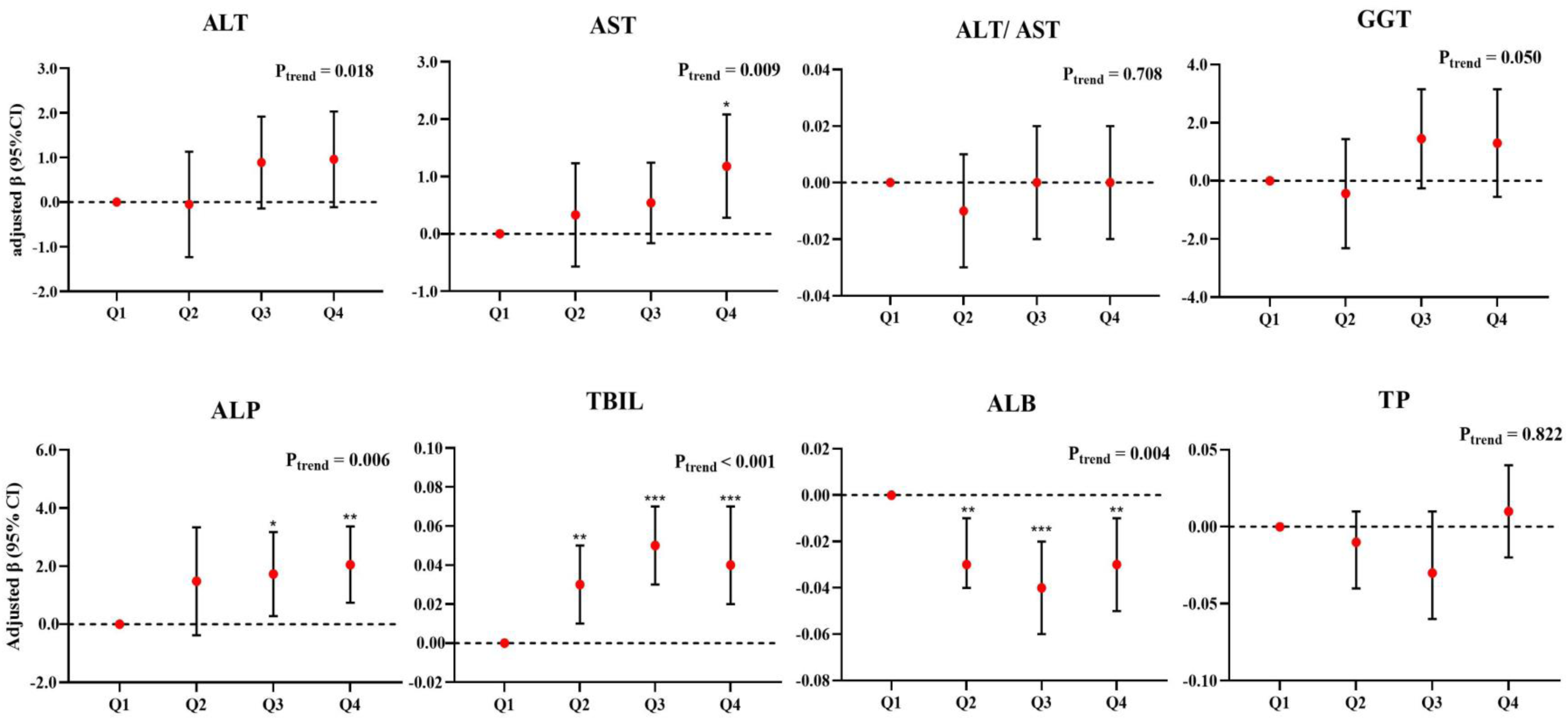
Adjusted associations of urinary BPA exposure with liver function parameters of all the participants, stratified by ln(BPA/Cr) quartiles. Model adjusted for age, sex, BMI, educational level, race/ethnicity, PIR, smoking status, alcohol consumption, hypertension, diabetes, and cardiovascular disease. \**P* <0.05, \*\**P* <0.01, \*\*\**P* <0.001

Stratified analyses showed that the highest quartile of urinary BPA was positively related to a higher level of serum ALT, AST, ALT/AST, GGT, and ALP in smokers, with statistical differences for ALT, ALT/AST, and GGT by smoking (z-test P< 0.05) (Figure 3). However, no difference was observed for TBIL and ALB by smoking (Figure 3). When stratified by alcohol drinking, a statistically higher level of ALT, AST, and GGT was found in drinkers after BPA exposure. However, there was no difference for other liver function indicators (z-test P > 0.05) (Figure 3). Figure S3 displays the associations by sex. Higher BPA exposure was related to elevated AST and TBIL in both men and women (Figure S3). Moreover, higher BPA exposure was positively associated with ALT and ALP in men, but with no sex difference (Figure S3). In the stratification by age, the association between BPA exposure with ALP was stronger in participants aged 60 years and older (Figure S4).

**Figure 3.**
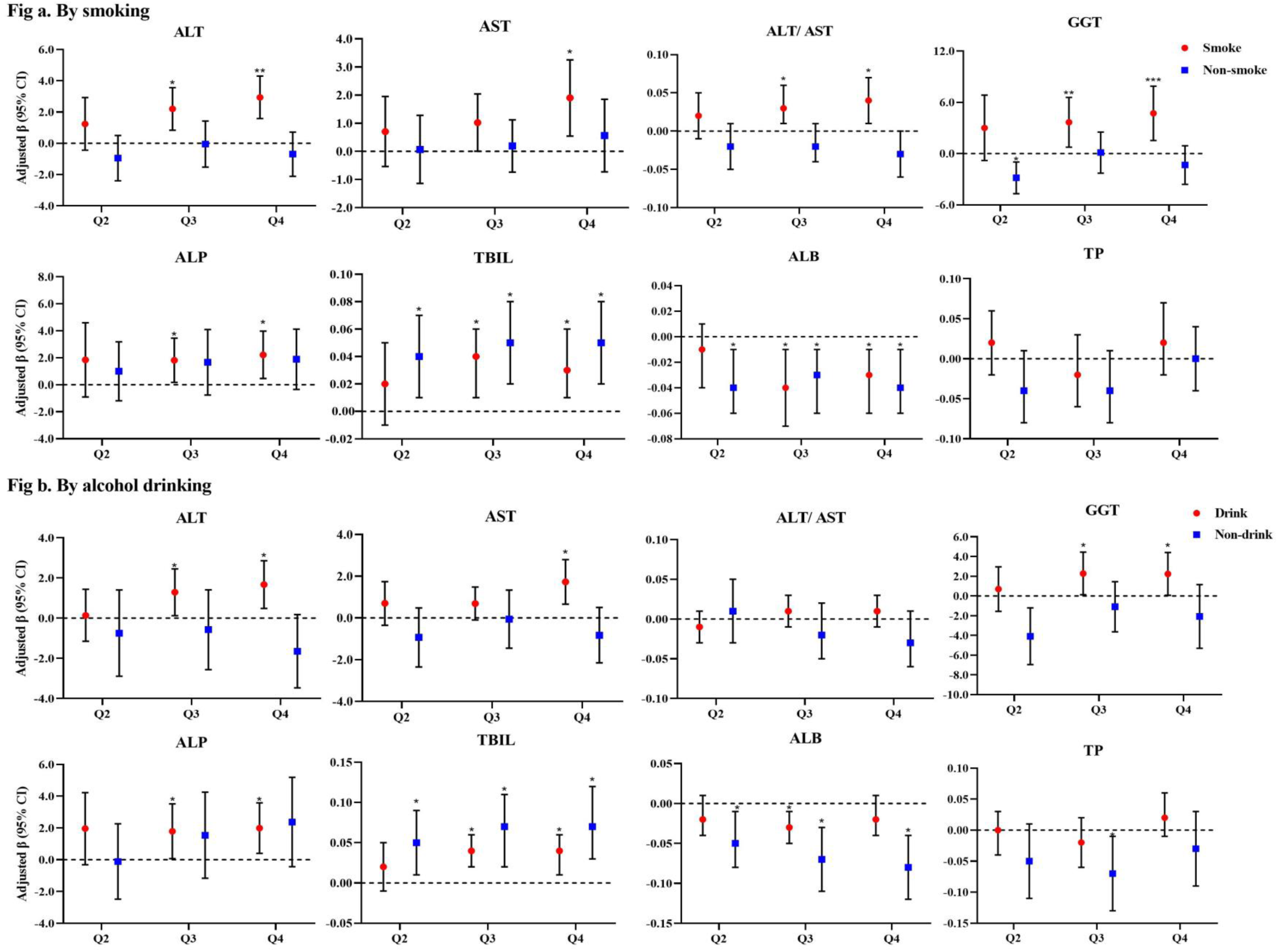
Adjusted associations of urinary BPA with changes in liver function indicators stratified by smoking or alcohol drinking. Note: Model adjusted for age, sex, BMI, educational level, race/ethnicity, PIR, smoking status, alcohol consumption, hypertension, diabetes, and cardiovascular disease except for the stratified variable. ALT: Alanine aminotransferase; AST: Aspartate aminotransferase; GGT: Gamma glutamyl transferase; AP: Alkaline phosphotase; TB: Total bilirubin; ALB: albumin; TP: Total protein. All the analyses conducted by using the lowest quartile as a reference. \**P* <0.05, \*\**P* <0.01, \*\*\**P* <0.001

## Discussion

Using a nationally population-representative sample from NHANES, our study explored the association between urinary BPA with eight liver function indicators among U.S. adults. The findings demonstrated that higher BPA exposure was related to elevated AST, ALP, and TBIL, while inversely with ALB. An exposure-response relationship was observed for urinary BPA with ALP and TBIL. The association for specific liver parameters (i.e., ALT, AST, GGT, or TBIL) was stronger in smokers or alcohol drinkers. Moreover, we found that individuals aged 60 years and older had a higher level of ALP after exposure to BPA.

The mean concentrations of urinary BPA in the present study ranged from 2.39 ng/ml to 5.57 ng/ml from 2003 to 2016, which was higher than that in Korea (mean 0.58 ng/ml) (16) and Canada (mean 1.16 ng/ml) (26). The major source of BPA is the production of polycarbonate (PC) plastics and epoxy resins, including food contact materials, protective linings for beverages and canned foods (27). According to a previous study from NHANES, the daily intake level of BPA was estimated to be 49.1-59.0 ng/kg/day for men and 36.2-46.5 ng/kg/day for women, respectively (28). To our knowledge, limited population-representative studies have explored the impacts of urinary BPA exposure on liver function indicators. Consistent with a previous study among U.S. adults, higher BPA concentration was associated with an elevated level of liver enzymes including GGT and ALP (17). A panel study conducted in Korea showed that higher BPA exposure was positively related to elevated AST, ALT, and GGT in the elderly, in line with our findings (16). Another study performed in Korean adults revealed that urinary BPA increased the prevalence of nonalcoholic fatty liver disease (NAFLD) and abnormal activity of ALT, AST, and GGT (29). Nevertheless, previous studies on these associations were based on limited liver function indicators. In the current study, we contributed to comprehensively assess the effects of urinary BPA on multiple liver indicators in a U.S. general population, not only including liver enzymes but also TBIL, ALB, and TP. In addition, we revealed the trend of urinary BPA concentrations across 14 years in seven cycles.

Our findings added the knowledge that higher BPA was related to elevated levels of AST, ALP, and TBIL, while inversely with ALB among U.S. adults.

The potential mechanisms of BPA exposure with abnormal liver function remain unclear. There are several possible explanations. One explanation is the direct toxicity of BPA to the liver. The liver plays an important role in metabolizing and detoxicating harmful chemicals (30), and these chemicals including BPA can accumulate in the liver and increase the vulnerability of liver dysfunction (16). Moreover, animal experiments in mice showed that low BPA dose could influence de novo fatty acid synthesis through increased expression of lipogenic genes, thus contributing to hepatic steatosis (31). Studies also demonstrated that oral BPA intake could induce oxidative stress in mice hepatocytes causing hepatocyte damage (32). Additionally, BPA exposure can result in the differential expression of genes related to lipid metabolism and adipogenesis and increasing 3T3-L1 adipocyte differentiation, which disturbs liver function (33,34).

In this study, we contributed to revealing the association of urinary BPA with specific liver function indicators (i.e., ALT, AST, and GGT) was stronger in smokers and alcohol drinkers in the U.S. A previous population-based study (35) showed that smoking was related to a higher level of serum ALT and ALP, similar with our findings. It is noted that tobacco smoke is a risk factor for liver disease, chemical compounds in tobacco have cytotoxic potential which increases necroinflammation and liver fibrosis. Moreover, smoking can increase the production of pro-inflammatory cytokines involved in liver cell injury (36). Regarding alcohol drinking, both epidemiological and toxicologic studies showed that alcohol consumption induces liver damage and leads to liver diseases through multiple pathways (37,38). Compared with other liver enzymes, serum GGT is involved in glutathione metabolism and is a well-established and more sensitive indicator for heavy alcohol drinking (39). In this study, we observed a higher GGT in drinkers, which might suggest a synergistic effect between BPA exposure with alcohol consumption. In addition, we found a higher ALP in participants aged 60 years and older with BPA exposure. Evidence has indicated that aging is related to significant loss of hepatic volume and blood flow, structural changes in liver cells, accumluation of aging pigments at the cytoplasm (40,41). However, there was no sex difference for BPA exposure with liver function indicators. Future studies in other populations are warranted to examine our findings.

Our study has several strengths. First, this was one of the few national-scale studies to evaluate the impacts of urinary BPA on a series of liver function parameters. The findings provide epidemiological evidence and insights for the prevention of liver injury. Second, based on a large population-representative sample from NHANES, we included seven cycles across 14 years, and the data revealed the trend of BPA exposure concentration in urine among the U.S. general population. Third, we built upon the knowledge to assess liver function indicators comprehensively rather than self-reported history of liver disease to reduce reporting bias. Last, the stratified analysis showed a stronger association with BPA exposure in smokers and alcohol drinkers, which highlighted that interventions are merited to protecting liver health for those vulnerable populations.

Several limitations should be acknowledged. First, since NHANES is a cross-sectional study, the causal relationship between urinary BPA with liver function indicators cannot be established. Second, considering the test of spot urine BPA only reflects recent BPA exposure, our study has the limitation of assessing the long-term effects of BPA. Moreover, some liver enzymes have short-term within-person variability (42) and one-time measurement may be not sufficient to identify the early stages of reversible liver damage (43). Third, our study used a representative U.S. population, additional studies with other populations may be necessary to generalize our findings. Last, although we collected much individual information in this study, the unobserved or residual confounding is inevitable due to the observational design.

## Conclusions

Using a nationally population-representative study, our findings identify the associations between urinary BPA exposure with multiple liver function indicators among U.S. adults. Higher BPA exposure was positively associated with AST, ALP, and TBIL, while inversely with ALB. The association for specific liver function indicators (i.e., ALT, AST, GGT, and TBIL) was stronger in smokers or alcohol drinkers. Our study provides important implications for policymakers to implement interventions to improve liver and planetary health.

## Data Availability

All data produced are available online at NHANES website.

## Author contributions

**Tiantian Zhang**: Investigation, Writing Writing-original draft, Obtaining funding. **Guang Huang**: Methodology, Software, Writing-review & editing. **Tongshuai Wang**: Writing review & editing. **Jie Chen**: Writing review & editing. **Xiangyu Zhou:** Writing review & editing. **Wenming Shi**: Investigation, Methodology, Acquisition, analysis, and interpretation of data, Writing review & editing, Supervision. All authors confirmed that they had full access to the study and accept responsibility to submit for publication.

## Conflict of interests

The authors declare that they have no competing interests.

## Funding

This work was supported by the National Natural Science Foundation of China (72204048).

## Abbreviations

BPA: Bisphenol A
NHANES: National Health and Nutrition Examination Survey
ALT: Alanine aminotransferase
AST: Aspartate aminotransferase
GGT: Gamma glutamyl transferase
ALP: Alkaline phosphotase
TBIL: Total bilirubin
ALB: Albumin
TP: Total protein
95% CI: 95% confidence interval
CVD: Cardiovascular disease
PIR: Poverty income ratio

## Notes

**Conflicts of interests** None declared.

### Competing Interest Statement

The authors have declared no competing interest.

### Author Declarations

The study was approved by the ethical review committee of the National Center for Health Statistics.

